# Brain Age Gap and Cognitive Processing Speed in Multiple Sclerosis

**DOI:** 10.64898/2026.08.20.26360954

**Authors:** Rodney Lea, Stasson Lea, Oun Al-Iedani, Vicki E. Maltby, Saad Ramadan, Jeannette Lechner-Scott

## Abstract

**Background and Objectives:** Cognitive impairment is common in multiple sclerosis (MS), but whether brain age gap (BAG) has greater cognitive relevance in MS than in people without brain disease is not known. We tested whether BAG was more strongly associated with cognitive processing speed (CPS) in MS.

**Methods:** We performed a cross-sectional analysis of MRI-derived BAG and CPS from a UK Biobank study consisting of 21,117 normative reference subjects with no recorded brain disease and 97 subjects with MS. BAG and CPS were standardized to the normative reference distribution, and an age- and sex-adjusted interaction tested whether the association differed between groups. Separately, a meta-analysis of the relationship of BAG and CPS was performed using published data from five independent MS cohorts (n=1,250 subjects in total). Correlation statistics were pooled to establish the effect size, 95% confidence intervals and p-values.

**Results:** In UK Biobank, there was a moderate negative association between BAG and CPS in MS (r=-0.35, 95% CI -0.52 to -0.17; P<.001), whereas the association in the normative reference group was negligible (r=-0.05, 95% CI -0.07 to -0.04; P<.001). There was a BAG-by-MS interaction indicating an MS-specific correlation (β =-0.19, 95% CI -0.29 to -0.09; P<.001). Across five independent clinical MS cohorts, the pooled BAG-CPS correlation was r=-0.25 (95% CI -0.33 to -0.18; P<.001). Overall, the magnitude of the association between BAG and CPS was at least five-fold greater in MS than in the normative population.

**Conclusion:** BAG was substantially more strongly associated with CPS in MS than in the normative population. These cross-sectional findings support further evaluation of BAG as an adjunctive MRI marker. Further studies are required to establish mechanism, prognosis, or clinical decision utility.

## Introduction

Cognitive impairment can occur early in multiple sclerosis (MS) and adversely affects employment and everyday function.^1,2^ Slowed information processing is among the most characteristic deficits. The Symbol Digit Modalities Test (SDMT) is primarily a measure of information-processing speed, although performance also depends on attention, visual scanning, and working memory. It is sensitive, reproducible, and practical for repeated assessment, and expert recommendations support routine cognitive screening in MS care.^3-5^ However, cognitive status is inconsistently assessed in clinical practice.

Brain-age modeling converts structural MRI into an estimate of apparent biological brain age relative to healthy aging patterns.^6^ Brain age gap (BAG), calculated as predicted minus chronological age, summarizes deviation from expected brain age. In MS, BAG has been associated with Expanded Disability Status Scale scores, lesion burden, brain atrophy, and subsequent disability accumulation.^7-10^ Estimates generally indicate that people with MS have brains appearing 4-6 years older than expected for their chronological age.^7,8,11-13^

The cognitive implications of this deviation are less clear. Higher BAG has been associated with poorer cognitive processing speed in MS.^11,12,14,15^ However, previous studies have generally estimated BAG-cognition associations within MS cohorts without testing whether BAG confers greater cognitive relevance in MS than in people without recorded brain disease. This BAG-cognition relationship may reflect a general association between older-appearing brain structure and cognition; alternatively, MS-related demyelination, diffuse white-matter injury, and gray-matter loss may modify the cognitive correlates of a given BAG.

We addressed this question using 2 complementary analyses. First, we tested whether the relationship between BAG and the Symbol Digit Substitution Test (SDST) differed between MS and a normative reference population.^13^ The UK Biobank SDST is an automated symbol digit processing speed task. SDST performance is strongly correlated with conventional SDMT performance, supporting its use as a comparable test of broad cognitive processing speed.^16^ Second, we conducted a targeted meta-analysis of 5 independent MS cohorts reporting BAG and clinical SDMT. We hypothesized that the association would be more negative in MS than in the normative reference population and reproducible across independent clinical cohorts.

## Methods

### Study Design

The study comprised an individual-level UK Biobank analysis and a targeted meta-analysis of published MS cohorts (Figure 1). The primary evidence for a group difference in the BAG-CPS association was the BAG-by-MS statistical interaction in the individual-level UK Biobank analysis, estimated using a common imaging and SDST platform.^13^ An independent clinical-SDMT meta-analysis assessed whether a negative BAG-CPS association was also observed across independent MS cohorts. Although correlated and comparable, SDST and SDMT estimates are not interchangeable and were not statistically pooled. For consistency, cognitive processing speed (CPS) is used hereafter as the umbrella term for this shared cognitive domain; SDST and SDMT are retained where the specific instrument or instrument-specific estimate is relevant.

**Figure 1.**
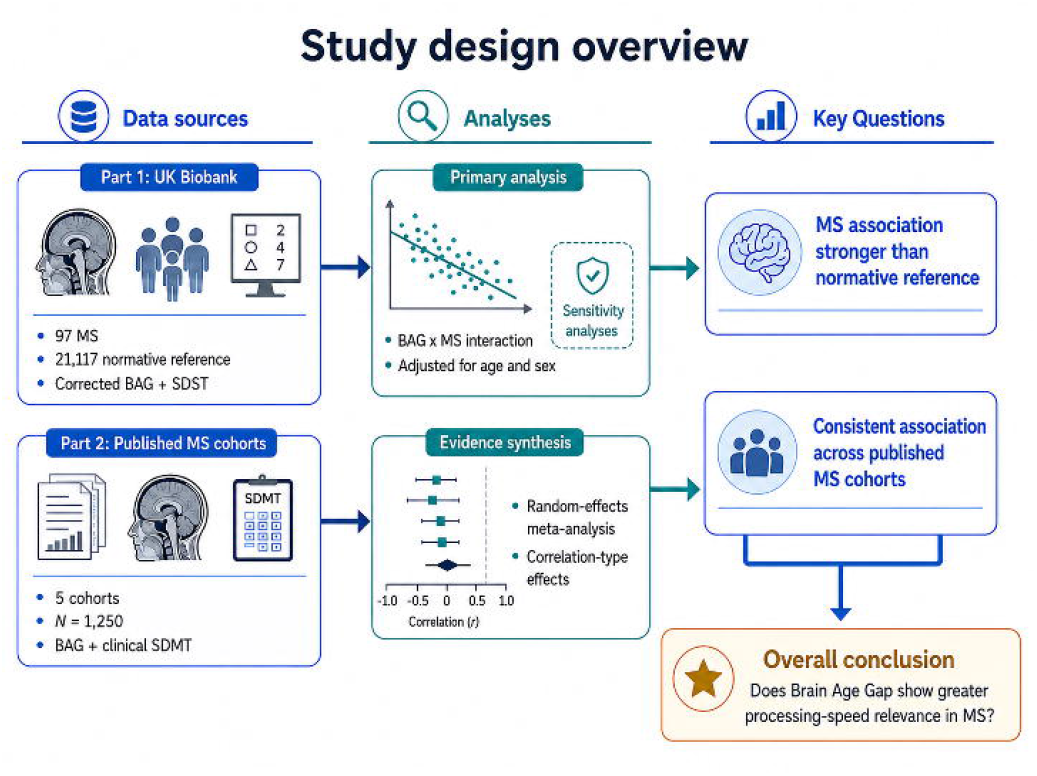
Study design overview. Part 1 used individual-level UK Biobank data to test whether the relationship between brain age gap (BAG) and Symbol Digit Substitution Test (SDST) performance differed between participants with multiple sclerosis (MS) and a normative reference population. Part 2 pooled correlation-type effects from 5 published MS cohorts reporting BAG and clinical Symbol Digit Modalities Test (SDMT). The diagram presents the study design and key questions rather than numerical results.

**Figure 2.**
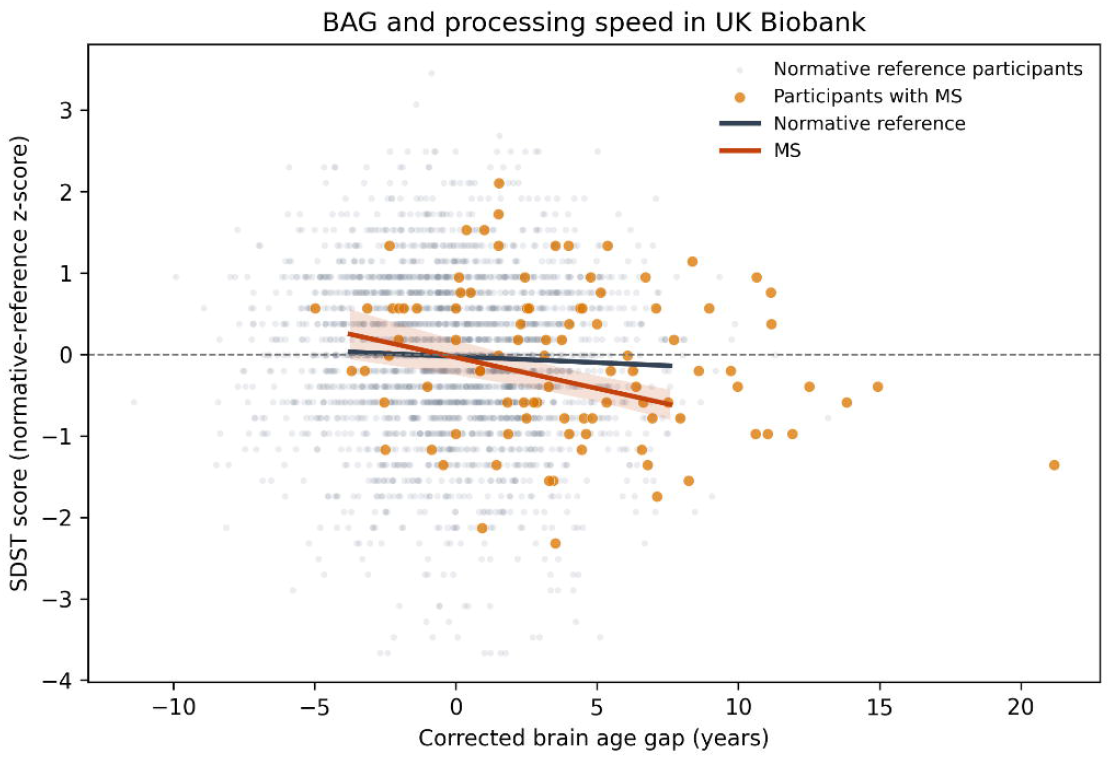
Relationship between corrected BAG and SDST performance in UK Biobank. All 97 participants with MS and a random sample of normative-reference participants are displayed; all 21,117 normative-reference participants contributed to model estimation. Lines and 95% confidence bands represent model-estimated relationships adjusted for age and sex. SDST was standardized to the normative-reference distribution.

### UK Biobank Data and Participants

Participant-level source data released with a published UK Biobank brain-age study were linked across cognition, diagnosis, and demographic files.^13^ The MS group comprised 97 participants identified as having multiple sclerosis in the source disease file (corresponding to ICD-10 code G35) who also had complete BAG, SDST, age, and sex data. The normative reference population comprised participants with complete data and no supplied diagnostic code for neurologic, neurodegenerative, cerebrovascular, or psychiatric disease; non-neurologic comorbidity was permitted. Predicted brain age and corrected BAG were obtained directly from the participant-level source data.^13^ Corrected BAG represented predicted brain age minus chronological age after correction for age-related prediction bias and remained expressed in years. Participant linkage and reference-population construction are detailed in the eMethods and eTable 2.

### Statistical Analysis of UK Biobank Data

Corrected BAG and SDST were standardized to the normative reference distribution. The primary linear model included standardized BAG, MS status, their interaction, age, and sex; coefficients were reported with 95% confidence intervals. The MS-specific BAG slope was obtained as the sum of the normative-reference BAG coefficient and the BAG-by-MS interaction. Group-specific age- and sex-adjusted partial correlations were estimated. Confidence intervals for these correlations, and for their difference, were derived from 5,000 stratified bootstrap replicates. Prespecified sensitivity analyses included 1:4 age- and sex-matching, overlap weighting, alternative covariate adjustment, flexible age modeling, influence-resistant estimation, and restriction to the overlapping BAG range. Analyses were conducted in Python 3.13.5 using NumPy 2.3.5, pandas 2.2.3, SciPy 1.17.0, statsmodels 0.14.6, and Matplotlib 3.10.8.

### Targeted Literature Search and Meta-analysis

A targeted structured search of PubMed/MEDLINE and Google Scholar was updated on July 28, 2026, without date restriction. Search terms combined MS with brain-age terminology and SDMT, cognition, or processing-speed terms; backward and forward citation chaining was also performed. Eligible reports were peer-reviewed full articles describing adults with MS, MRI-derived BAG or an equivalent measure, contemporaneous clinical SDMT, and sufficient information to derive a signed correlation-type effect and sampling variance. Duplicate cohorts, inseparable non-MS effects, non-SDMT outcomes, longitudinal-only effects, and preprints were excluded from the primary synthesis. Search strings, screening rules, and full-text exclusions are reported in the eMethods and eTables 3 and 13 in the eSupplement.

Five independent cohorts from 4 peer-reviewed studies were included.^11,12,14,15^ Adjusted partial correlations, Pearson correlations, Spearman correlations, and recoverable regression results were transformed to Fisher z. Estimates were pooled using inverse-variance restricted maximum-likelihood random effects with modified Knapp-Hartung inference and back-transformed to r.he studies differed in brain-age algorithm and age-bias correction; these characteristics were retained as study-level descriptors and examined in sensitivity analyses rather than assumed to be equivalent. Sensitivity analyses examined uncertain effect reconstructions, alternative BAG processing, and leave-one-cohort-out results. Detailed derivations are provided in eTables 6-9.

### Data Availability

Deidentified participant-level source data used for the UK Biobank analysis were released with Zhang et al.^13^ and remain subject to the source data-use conditions. Derived analysis datasets, study-level meta-analysis inputs, analysis code, and reproducibility materials will be made available through GitHub at [GITHUB REPOSITORY URL TO BE INSERTED].

### Standard Protocol Approvals, Registrations, and Patient Consents

The present study used only deidentified, publicly released participant-level data and published aggregate results; therefore, no additional local ethics review or participant consent was required. The source UK Biobank study was conducted under North West Multi-centre Research Ethics Committee approval (06/MRE08/65), and all participants provided written informed consent.^13^

## Results

### UK Biobank Population

The analytic sample included 21,117 participants in the normative reference population (mean age 64.6 years; 51.3% female) and 97 participants with MS (mean age 60.9 years; 77.3% female). Mean corrected BAG was 0.06 years in the normative reference population and 3.90 years in MS; mean unadjusted SDST scores were 19.06 and 18.59, respectively (eTable 1).

### BAG-CPS Association by MS Status

Among participants with MS, higher BAG was moderately associated with poorer CPS (r=-0.35, 95% CI -0.52 to -0.17; P<.001). In contrast, the association in the normative reference population was negligible (r=-0.05, 95% CI -0.07 to -0.04; P<.001). The stronger association in MS was supported by the BAG-by-MS interaction coefficient of -0.19 (95% CI -0.29 to -0.09; P<.001), with a bootstrap difference in partial correlations of -0.30 (95% CI -0.46 to -0.12; eTable 11). On the raw scale, each additional BAG year corresponded to 0.40 fewer SDST points in MS compared with 0.08 fewer points in the normative reference population. The association remained in the same direction after making the groups more comparable in age and sex. Full sensitivity results are provided in eTables 4 and 5 and eFigures 1 and 4.

### Meta-analysis of Published MS Cohorts

The 5 independent cohorts from 4 studies comprised 1,250 participants with MS.^11,12,14,15^ All cohort-specific estimates were negative despite differences in brain-age algorithm, BAG correction, and SDMT administration (Figure 3; eTables 6 and 7). The pooled correlation indicated a moderate association between higher BAG and poorer CPS measured by clinical SDMT (r=-0.25, 95% CI -0.33 to -0.18; *P*<.001). Estimated between-cohort heterogeneity was zero (I^2^=0%), but with only 5 cohorts the analysis had limited ability to detect or explain between-study variation. Pooled estimates ranged from -0.26 to -0.24 across most sensitivity analyses (eTables 8 and 9; eFigure 2).

**Figure 3.**
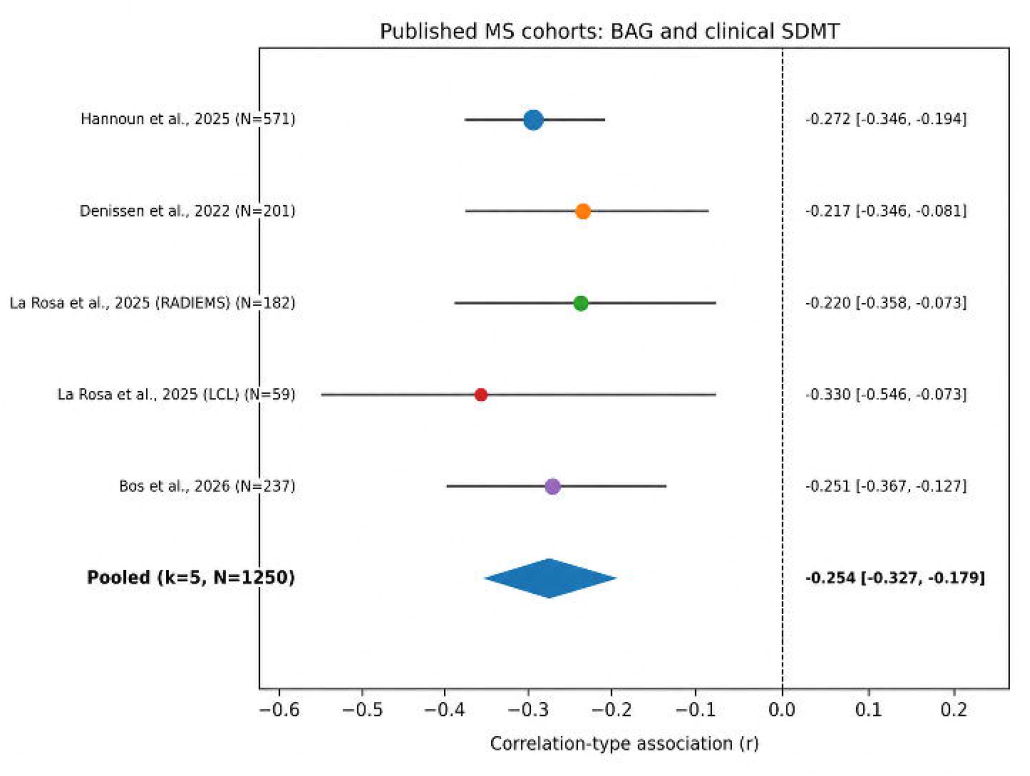
Forest plot of 5 independent published MS cohorts. Correlation-type effects were pooled on the Fisher z scale using restricted maximum-likelihood random effects and modified Knapp-Hartung inference. Circles show cohort estimates with 95% CIs; the diamond shows the pooled estimate. SDMT = Symbol Digit Modalities Test.

### Integrated Findings

The UK Biobank MS estimate (r=-0.35) was concordant in direction and broadly similar in magnitude to the pooled clinical-SDMT estimate (r=-0.25), supporting consistency across independent datasets and related measures of CPS. Because the UK Biobank analysis used SDST and the clinical meta-analysis used SDMT, which are correlated but nonidentical instruments, the estimates were interpreted as complementary evidence and were not statistically pooled. Overall, the magnitude of the association between BAG and CPS was at least five-fold greater in MS than in the normative population (Figure 4, eFigure 3).

**Figure 4.**
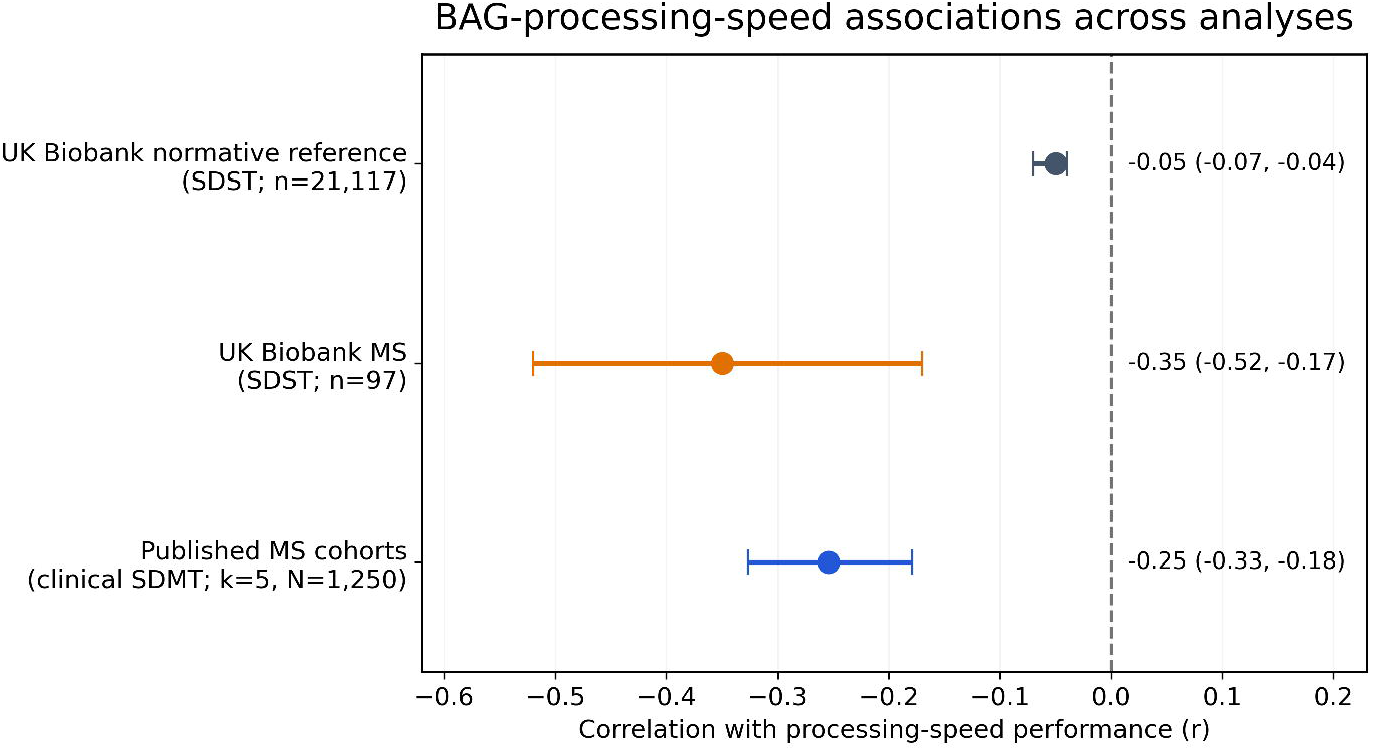
BAG-CPS associations across the normative reference and MS analyses. UK Biobank estimates are age- and sex-adjusted partial correlations using SDST; the published-cohort estimate is the pooled correlation-type effect using clinical SDMT. The difference in partial correlations between the UK Biobank MS and normative-reference groups was -0.30 (bootstrap 95% CI -0.46 to -0.12). UK Biobank SDST and conventional SDMT are related but nonidentical CPS measures. Their estimates are displayed together for interpretation and were not combined in a pooled model.

## Discussion

Taken together, the UK Biobank and independent clinical-cohort analyses indicate that BAG has much greater correlation to CPS in MS than in the normative population, with consistent effects across independent MS cohorts. BAG elevation in MS and its association with CPS reflect distinct phenomena. Previous studies have established that people with MS have brains appearing 4-6 years older than expected for age.^7,8,11-13^ The present findings extend this literature by indicating that the cognitive relevance of a given BAG differs by disease context rather than merely reflecting greater BAG values in MS.

Brain age estimates reflect general structural features of the brain that include gray and white matter loss and ventricular expansion. In MS, these features coexist with focal lesions, demyelination, axonal injury, and network disruption.^2,17^ One possible interpretation is that a given increase in BAG reflects a structurally more consequential disease context in MS than in participants without recorded brain disease. This interpretation is consistent with prior associations between BAG, lesion burden, atrophy, disability, and progression.^7-12^ The present analysis did not measure neural reserve or establish this mechanism directly, so this interpretation remains a hypothesis.

The observed association was moderate and does not establish diagnostic or prognostic utility. BAG may ultimately provide complementary structural information when direct cognitive assessment is unavailable or has not been performed, but its incremental clinical value over SDMT, conventional MRI, and clinical variables remains to be established. In future clinical workflows, an unexpectedly high BAG could strengthen the rationale for cognitive screening, repeat assessment, or neuropsychological referral when cognition has not otherwise been evaluated.

BAG should be interpreted alongside age, mood, fatigue, Expanded Disability Status Scale score, lesion burden, atrophy, and direct cognitive assessment. Its potential value lies in adding an intuitive global MRI summary to existing information, not in functioning as a stand-alone diagnostic test.

Key strengths were the large normative reference group, use of a common imaging and SDST platform for the UK Biobank comparison, and independent supporting evidence from 5 published MS cohorts using different brain-age methods. The UK Biobank and clinical-SDMT analyses were kept separate because SDST and SDMT are related but nonidentical instruments. This study had several limitations. First, only 97 UK Biobank participants with MS had complete BAG and SDST data, limiting subgroup and nonlinear analyses. Second, the normative reference population was defined through exclusion of supplied coded brain disorders rather than comprehensive clinical screening. Third, although UK Biobank SDST and conventional SDMT show substantial convergent validity and assess the same broad symbol-digit processing-speed domain, they are nonidentical instruments. Their results were therefore interpreted as complementary and were not combined in a single pooled estimate. Fourth, all analyses were cross-sectional and do not establish causality, prediction of future cognitive decline, or clinical decision utility.

Overall, BAG was substantially more strongly associated with CPS in UK Biobank participants with MS than in the normative reference population, and the separate clinical-SDMT synthesis showed a concordant moderate association. These findings support further evaluation of BAG as an adjunctive MRI marker associated with cognitive performance, but they do not establish mechanism, prognosis, or clinical decision utility.

## Data Availability

All data produced in the present study are available upon request to the authors

## Acknowledgments

The authors acknowledge the participants that consented and contributed data used in this study. OpenAI ChatGPT (GPT-5.6 Pro; OpenAI, 2026), Anthropic Claude [Sonnet5, Anthropic, 2026], and Google Gemini Pro (3.6 Flash, Google, 2026) were used to assist literature search, statistic extraction and cross-checking, manuscript organization, and language editing. Rodney Lea reviewed and verified all AI-assisted outputs against the source data and original publications, checked the references and numerical results, and approved the final text. The scientific interpretation and conclusions are those of the authors.

